# Beyond word error rate: clinical risk as the necessary standard for ambient AI scribe evaluation: evidence from 77 global languages

**DOI:** 10.64898/2026.09.10.26362715

**Authors:** Henry Bergman, Vivian Liu, Ben Austin, Rohan Sanghera

## Abstract

**Objective:** Ambient AI scribes evaluated using frequency-based metrics such as word error rate (WER), which do not represent clinical consequence. We tested whether variation in these metrics tracks consequential transcription errors.

**Methods:** We constructed a multilingual corpus from five clinical dictation scripts spanning a complexity gradient, translated into 99 languages, rendered to synthetic speech under three acoustic conditions, and transcribed by a production ambient scribe. Six frequency metrics were computed. Three independent large language model raters from external providers assessed clinically meaningful error patterns in context using a Severity x Likelihood framework informed by UK digital clinical-safety-risk-management principles.

**Results:** Across 59,819 genuine transcription-error occurrences, 58,329 (97.5%) were LOW risk and 251 (0.42%) CRITICAL or HIGH. None of six frequency metrics showed a statistically detectable association with serious clinical risk across languages; correlations were small (absolute Spearman ρ<0.16). A Severity × Likelihood sum remained strongly correlated with WER (ρ=0.80), showing that the aggregate remained dominated by benign errors. At complexity level 3, low-resource languages had worse WER than high-resource languages (β=+0.078, 95% CI +0.045 to +0.111; p<0.0001), without a detectable difference in CRITICAL/HIGH risk (OR 1.21, 95% CI 0.43 to 3.43; p=0.72).

Consultation complexity was the principal predictor of serious risk (OR 3.06 per level, p<0.0001).

**Conclusion:** Across this controlled multilingual corpus, aggregate transcription-frequency metrics did not reliably track the sparse severe tail of clinically consequential errors. WER remains appropriate for transcription quality, but these data do not support its use alone as a proxy for clinical safety.

Context-aware assessment of error consequence provides complementary information that frequency measures can dilute.

**Highlights:**

- WER did not track serious clinical-risks rate across 77 languages
- Five related frequency metrics showed the same dissociation
- Severity-weighting remained dominated by common low-risk errors
- Quality tracked language resource; serious risk tracked complexity
- We provide a reusable context-aware clinical-risk instrument

## 1. Introduction

Ambient artificial-intelligence scribes, which listen to a clinical encounter and produce a structured note, are among the fastest-deploying clinical AI applications, and are increasingly being adopted at scale across health systems and languages, with the explicit promise of reducing documentation burden while maintaining note quality [1,2]. As deployment globalises, the question of how these systems should be evaluated safely and at scale becomes increasingly consequential.

The de facto standard is word error rate (WER) and its relatives. WER measures the proportion of reference words substituted, deleted or inserted in a transcript; lexical- and embedding-based metrics including BLEU, ROUGE, chrF and BERTScore extend related overlap-based approaches to generated text. A systematic review found evaluations of AI-generated clinical notes to be dominated by lexical-overlap metrics [3]. These are fundamentally frequency-based measures: errors contribute according to their occurrence, without regard to clinical consequence. When used to support claims about safety, they embed a strong assumption that clinical risk rises with aggregate error frequency.

There is little reason to assume that relationship holds. A deletion may remove conversational filler or a drug allergy; a substitution may exchange synonyms or two anticoagulants. A single inverted negation or medication substitution can carry greater clinical consequences than hundreds of cosmetic mishearings. If risk is concentrated in a sparse severe tail that is only weakly coupled to overall error frequency, frequency metrics cannot reliably proxy safety.

This can be framed as a surrogate-endpoint problem. When a transcription metric is used to support inference about patient safety, it functions as a candidate surrogate for clinical harm. Surrogate validity requires an empirical relationship with the outcome represented [4], while medical-device risk-management principles assess hazards through both severity and likelihood [5-7]. We therefore test a necessary, rather than sufficient, condition for surrogate use: whether variation in commonly used frequency metrics is associated with variation in directly measured clinical risk.

The clinical speech-recognition literature exposes the same measurement gap. Two systematic reviews found weak and heterogeneous evidence, inconsistent definitions and wide variation in reported WER and evaluation practice [8,9]. Neither established the clinical consequence of individual errors as an outcome distinct from aggregate frequency. Among 217 dictated notes, 15.8% of raw speech-recognition errors involved clinical information and 5.7% were clinically significant [10]; clinician surveys and editing studies further suggest that raw transcription accuracy incompletely represents what reaches the record [11,12]. In generated clinical text, lexical-overlap metrics also penalise meaning-preserving paraphrases [3], prompting dedicated clinical-safety frameworks and expert-panel assessment [13,14].

The methodological caution is familiar from surrogate-endpoint research: mechanistic plausibility or association alone is insufficient to establish validity [15]. We use this literature as a validity framework without claiming complete Prentice validation; patient-safety research also shows that observed event rates depend on detection sensitivity [16]. Automated speech recognition performs unequally across populations and languages [17,18], while language discordance is associated with adverse events and incomplete communication [19,20]. Medication-name confusion and negation or assertion-status errors are plausible severe-tail mechanisms: look-alike and sound-alike medicines remain recognised sources of medication error [21-23], and negation is a longstanding clinicallanguage-processing failure mode [24].

We tested the specific, falsifiable hypothesis that aggregate frequency metrics are dissociated from clinical risk. If WER is informative about safety, languages with poorer WER should also exhibit more clinically dangerous errors and similar rankings by accuracy and risk; if consequential errors occupy a sparse severe tail weakly related to overall frequency, those rankings should diverge. Across six metrics and 77 languages, we test this relationship, introduce a reusable three-rater context-aware risk instrument, and examine whether the established accuracy disadvantage in lower-resource languages is accompanied by a clinical-risk gradient. The study deliberately isolates the transcription layer: subsequent note generation may correct, propagate or amplify transcription errors and introduce errors independently, so transcription-layer safety cannot itself establish the safety of the clinician-facing note.

## 2. Methods

### 2.1 Design and rationale

We compared two constructs: transcription-error frequency and context-aware clinical risk. A fully specified synthetic corpus provided an exact reference, controlled variation and between-language heterogeneity. This strengthened internal validity for metric comparison while limiting inference about real-world multilingual performance, particularly for equity analyses.

### 2.2 Evaluation corpus and complexity gradient

Five English clinical dictation scripts formed a deliberate complexity gradient: a simple primary-care consultation (L1); a moderate primary-care review with vital signs and lipids (L2); an outpatient diabetes review with medications and laboratory values (L3); a dense inpatient respiratory ward round (L4); and an expert oncology handover covering staging, molecular markers and chemotherapy (L5). Scripts were translated into 99 languages by a large language model, rendered in native script, checked by back-translation and frozen. Translation held clinical content as constant as practicable but may under-represent idiomatic or culturally specific language.

### 2.3 Speech synthesis and the neural-voice inclusion criterion

References were rendered using the highest-quality text-to-speech available, with neural multispeaker voices and two voices per language where possible. Languages limited to single-speaker non-neural synthesis were excluded from primary risk inference. Four unspaced neural-voiced languages were analysed separately using character-level alignment; Chinese and Japanese were not risk scored. The final corpus comprised 99 languages, 77 in primary risk inference.

### 2.4 Acoustic conditions and transcription

Each utterance was presented as clean audio, simulated clinic ambience and simulated ward noise, then transcribed through real-time capture by a production ambient scribe.

### 2.5 Frequency metrics

We computed script-aware WER, native-script medical entity error rate (ME-WER), BLEU, chrF, ROUGE-L and multilingual BERTScore. Normalisation addressed Unicode, punctuation, transliteration and unspaced scripts. ME-WER covered abbreviations, drugs, laboratory terms and proper nouns, providing the frequency measure most plausibly related to clinical risk [3].

### 2.6 Identifying genuine clinical errors

Before risk scoring, we removed semantically equivalent number formats, register or inflection variants, synonyms and spelling variants, and equivalent code-switching. Differences involving drugs, tests, values, doses, anatomy, diagnoses or negation were always retained. The procedure conservatively favoured retention of potentially clinical errors.

### 2.7 Clinical-risk instrument

Three large language models from different external providers and model families independently scored each genuine error pattern using a fixed rubric; none belonged to the transcription system. Severity (1-5) represented potential consequence if the error reached the record and was believed, and Likelihood (1-5) the probability of surviving review and propagating to harm. Raters viewed 110 characters of reference and hypothesis on each side of the error, so a value restated correctly nearby or an implausible garble received a lower propagation likelihood than the same surface error judged alone. Severity × Likelihood defined CRITICAL (16-25), HIGH (10-15), MEDIUM (5-9) and LOW (1-4); majority vote determined the consensus tier. The framework was informed by ISO 14971, DCB0129/DCB0160 and MHRA guidance [5-7]. Supplementary Files S1 and S2 provide the complete specification and method handover.

### 2.8 Resource classification and covariates

Languages were grouped by the Joshi resource taxonomy as high (classes 4-5), medium (class 3) or low (classes 0-2) [25]. Secondary covariates were GDP per capita of the dominant first-language country [26], first-language speaker availability [27] and language family.

### 2.9 Statistical analysis

Language was the unit of cross-language surrogate analysis. Observation-level models used 5,308 observations from 77 spaced, neural-voiced languages. We correlated each metric with the proportion of observations containing at least one CRITICAL/HIGH error using Spearman coefficients and bootstrap 95% confidence intervals; risk was also expressed per 1,000 reference words. A continuous Severity × Likelihood sum tested whether weighting all errors reconstructed frequency. A Gaussian mixed-effects model for semantic-normalised WER included a language random intercept; a cluster-robust logistic model examined serious risk. Both included resource level, complexity, noise and a resource-by-complexity interaction. Because each script represented one complexity level, script and complexity effects were not separately identifiable. Complexity was re-centred at level 3 for interpretable resource contrasts (Table S8). English contributed 46.1% of observations; English-excluded and volume-balanced analyses assessed this imbalance. Pairwise resource comparisons used Mann-Whitney U tests with Benjamini-Hochberg correction [28]. Prespecified subgroups covered complexity, noise and language family. We quantified reliability and simulated correlation power under the realised design: with 77 languages, median 30 observations each and a pooled serious-event rate of 5.31%, power was approximately 0.44 for a true Spearman correlation of 0.27; 80% power required approximately 0.40 (Analysis A1). Analyses used Python 3.9 and statsmodels 0.14.6.

## 3. Results

We analysed 5,758 observations across 99 languages. Clinical-risk inference was conducted on the 77 spaced, neural-voiced languages meeting the pre-specified inclusion criterion. Four unspaced neural-voiced languages (Burmese, Thai, Khmer, Lao) were analysed separately at the pattern level as a sensitivity extension. Unspaced Neural-voiced Chinese and Japanese were not risk scored. 16 languages with only single-speaker robotic synthesis were excluded from inference against our prespecified exclusion criteria. Frequency metrics are reported for all 99 languages.

### 3.1 Clinical risk is rare and tightly concentrated in a severe tail

Across 59,819 scored genuine-error occurrences drawn from 9,110 unique error patterns, 58,329 (97.5%) were LOW and 1,239 (2.1%) MEDIUM risk, while 191 (0.3%) were HIGH and 60 (0.1%) CRITICAL. CRITICAL and HIGH together accounted for 251 occurrences (0.42%) in the three-rater majority consensus (Table S2). Agreement among the automated raters was high in absolute terms: 93% of patterns received the same tier from all three raters and every pattern was resolved by majority. Unweighted Fleiss kappa was 0.358; because kappa is sensitive to highly skewed marginals, it is reported with raw agreement [30,31]. Quadratic-weighted and binary CRITICAL/HIGH kappa were each 0.52 [31,32], indicating predominantly adjacent-tier disagreement. The unspacedlanguage extension had a similar distribution (Table S7).

### 3.2 Frequency metrics did not satisfy the tested surrogate condition

No frequency metric showed a statistically detectable association with the rate of clinically dangerous errors across languages (Fig. 1). Spearman correlations between the proportion of observations containing a CRITICAL or HIGH error and each metric were near zero and nonsignificant: WER ρ=-0.16 (95% CI -0.37 to +0.07), ME-WER ρ=+0.12, BLEU ρ=+0.11, chrF ρ=+0.12, ROUGE-L ρ=-0.02 and BERTScore ρ=+0.03 (all p>0.05). The observed WER estimate therefore did not support a moderate positive association in the measured language-level rates. However, outcome-sparsity analysis showed reliability and power (Supplementary Analysis A1), so these results indicate failure to detect an association in this design rather than exclusion of smaller latent associations.

Concordant findings across differently constructed metrics, including clinically restricted ME-WER, were inconsistent with aggregate accuracy reliably identifying higher clinical risk.

### 3.3 Why the measures diverge: risk lives in a severe tail

When all errors were weighted by Severity × Likelihood and summed, the resulting score correlated strongly with WER (ρ=+0.80) and inversely with BLEU (ρ=-0.75). Common LOW-risk errors dominated the aggregate; divergence became visible only in the severe tail. Severity weighting alone therefore did not produce a clinically discriminating metric when the underlying error distribution was highly skewed.

### 3.4 Quality and risk are governed by different variables

Transcription quality and clinical risk responded to different predictors (Table 1). In the linear mixedeffects model, WER was statistically significantly worse for low-resource languages (beta = +0.068 versus high-resource, p < 0.001) and rose with complexity (β = +0.007 per level, p < 0.001) and with ward noise (β = +0.014, p < 0.001). Because the interaction model’s uncentred resource coefficients refer to complexity=0, outside the observed range, we re-centred complexity at level 3. At that level, low-resource languages had higher WER than high-resource languages (β=+0.078, 95% CI +0.045 to +0.111; p<0.0001), while their CRITICAL/HIGH odds did not differ detectably (OR 1.21, 95% CI 0.43 to 3.43; p=0.72; Table S8). Complexity was the principal detected predictor of serious risk (OR 3.06 per level, 95% CI 2.17 to 4.32; p<0.0001). Noise and the resource-by-complexity interaction were not statistically significant for serious risk. We therefore describe an absence of a detectable resource gradient rather than evidence of equivalence.

**Table 1.** Transcription quality and clinical risk are governed by different variables.

| Predictor | Effect on WER (quality) | Effect on CRITICAL/HIGH (risk) |
| --- | --- | --- |
| Language resource (low vs high) | $\beta = +0.068$ (95% CI +0.032 to +0.104), $p < 0.001$ | OR 3.15 (95% CI 0.32 to 31.06), $p = 0.33$ |
| Language resource (medium vs high) | $\beta = -0.027$ (95% CI -0.067 to +0.014), $p = 0.19$ | OR 0.84 (95% CI 0.03 to 20.13), $p = 0.91$ |
| Clinical complexity (per level) | $\beta = +0.007$ (95% CI +0.004 to +0.009), $p < 0.0001$ | OR 3.06 (95% CI 2.17 to 4.32), $p < 0.0001$ |
| Clinic noise (vs clean) | $\beta = +0.004$ (95% CI -0.003 to +0.011), $p = 0.30$ | OR 0.88 (95% CI 0.71 to 1.08), $p = 0.22$ |
| Ward noise (vs clean) | $\beta = +0.014$ (95% CI +0.007 to +0.021), $p < 0.0001$ | OR 1.20 (95% CI 0.94 to 1.54), $p = 0.14$ |
| Resource (low) × complexity | $\beta = +0.003$ (95% CI -0.002 to +0.008), $p = 0.19$ | OR 0.73 (95% CI 0.46 to 1.16), $p = 0.18$ |
| Resource (medium) × complexity | $\beta = +0.012$ (95% CI +0.006 to +0.018), $p < 0.001$ | OR 0.85 (95% CI 0.42 to 1.71), $p = 0.64$ |

### 3.5 Clinical risk increased principally with consultation complexity

No CRITICAL or HIGH errors were observed at L1 or L2. The proportion of observations containing at least one serious error rose to 3.4% at L3 and 11.6% across L4-L5 (Fig. 2), peaking at L4 in all resource strata rather than increasing monotonically. At L4, language-averaged CRITICAL/HIGH densities were 0.52, 0.17 and 0.49 per 1,000 words in high-, medium- and low-resource languages. FDR-corrected comparisons showed worse WER in low-resource than high- and medium-resource languages (FDR=0.001 and 0.009), while no serious-risk comparison survived correction (all FDR>0.30).

Language family explained additional WER variance (ΔR^2^=0.05, p<0.0001) but without additional serious-risk variation.

### 3.6 Secondary analysis

accuracy disparities were not accompanied by a detectable safety gradient Among 74 languages with complete external covariates, GDP per capita and first-language speaker availability correlated with WER (ρ=-0.25, p=0.031; ρ=-0.24, p=0.037), as did resource class among 77 languages (ρ=-0.46, p<0.001), but none correlated detectably with clinical risk (ρ=+0.03, +0.13 and +0.09; Table 2). The neural-versus-robotic voice comparison also supported the prespecified inclusion criterion: for languages with both voice types available, switching to a neural voice on the same transcriber reduced WER by 0.10 to 0.21. Further detail is reported in Tables S1, S3 and S4

### 3.7 The serious-error register is small and patterned

The consensus CRITICAL/HIGH set comprised 34 distinct error patterns, 9 CRITICAL and 25 HIGH, dominated by medication-name confusions and negation or clinical-status flips (Table 3; Table S5-S6). These error classes produce fluent and clinically plausible output, characteristics that may make them less conspicuous during review. Their concentration in a small number of recurring mechanisms suggest medication-specific confidence checks and assertion-status safeguards as testable mitigations, without demonstrating reduced downstream harm.

**Table 2.** Development proxies were associated with transcription quality but not detectably with serious clinical risk.

| Development proxy | Spearman $\rho$ vs WER (quality) | Spearman $\rho$ vs CRITICAL/HIGH (risk) |
| --- | --- | --- |
| GDP per capita | -0.25, $p = 0.031$ | +0.03 (ns) |
| First-language speaker availability | -0.24, $p = 0.037$ | +0.13 (ns) |
| Joshi resource class | -0.46, $p < 0.001$ | +0.09 (ns) |

Negative correlations against WER indicate that higher values of the proxy are associated with lower (better) word error rate. ns, not significant.

**Table 3.**
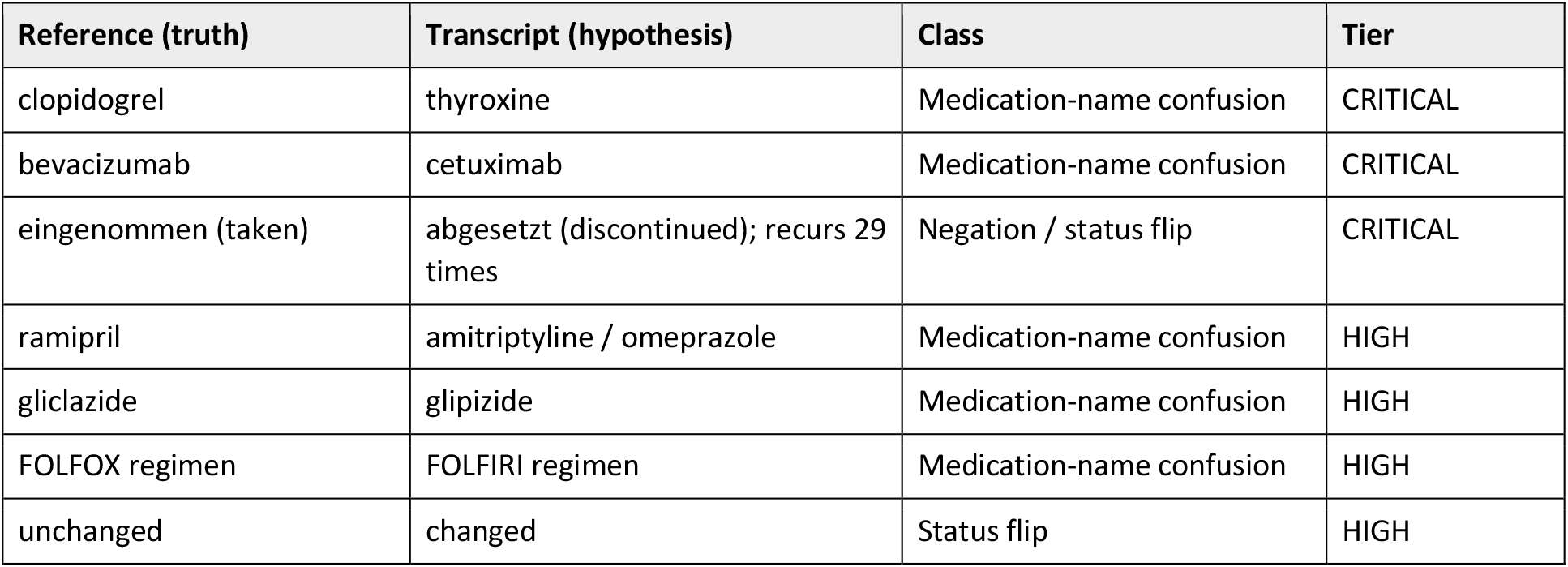
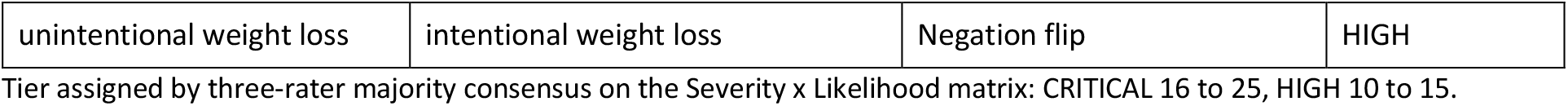
Serious-error register: representative CRITICAL and HIGH patterns.

## 4. Discussion

Across 77 languages and six metrics, aggregate transcription accuracy did not reliably identify languages with higher rates of clinically dangerous transcription errors. Associations between the frequency metrics and the CRITICAL/HIGH outcome were small and non-significant, while transcription quality and serious risk showed different predictor profiles: language resource and acoustics for WER, and consultation complexity for risk. Under the surrogate-endpoint framing, this failure to observe a necessary empirical relationship does not constitute complete Prentice validation [4,15].

The practical implication is narrower and more defensible than declaring WER invalid for its intended purpose. WER remains an appropriate measure of transcription accuracy and the frequency of errors. The present data do not support extending it, without further evidence, to a stand-alone claim about clinical safety. A frequency metric may still be useful for engineering quality control, but safety inference requires direct measurement of clinically consequential error or independent evidence that the chosen metric predicts such error in the target setting.

### 5.1 Why frequency and risk come apart

Clinical risk is heavy-tailed and concentrated in a small minority of errors, primarily medication-name substitutions and negation or status flips, whereas frequency metrics are dominated by the benign majority. The same problem persists when all errors are multiplied by severity and summed, because the mass of low-risk events still governs the aggregate. Capturing the safety-relevant signal therefore requires isolating the severe tail and judging errors in context.

A related hypothesis is that fluency may be double-edged. Rare errors produced by an otherwise accurate system can be difficult to detect precisely because the surrounding output is plausible, which is consistent with literature on automation bias [33] and evidence that many speechrecognition errors are corrected before signature [10], but our study did not directly measure clinician detection or downstream propagation and cannot establish this mechanism.

### 4.2 Implications for evaluation and deployment

Ambient-scribe evaluations should report error consequence alongside WER, prespecify safety-relevant outcomes and power analyses for those outcomes directly. Studies powered on aggregate error frequency may remain underpowered for consequential differences when serious errors are sparse.

Two operational hypotheses follow from these data. First, safety evaluation should be stratified by encounter complexity, because serious errors clustered in denser clinical scenarios. Second, the dangerous-error register suggests that medication-name confusion and assertion-status errors are high-yield targets for mitigation. These observations identify testable engineering priorities rather than demonstrated reductions in downstream patient harm.

### 5.4 Equity: no detectable safety gap despite a large accuracy gap

Low-resource languages and languages associated with lower-GDP or smaller-speaker-population countries had substantially worse transcription quality. This is an important inequity for documentation quality, clinician workload and trust, and is consistent with known disparities in speech technology [16,17]. In these data the quality gap did not produce a statistically detectable safety gap; however this finding should be interpreted cautiously: only 251 serious occurrences were observed, the low-resource odds-ratio estimate was clinically non-trivial but imprecise. Synthetic speech also does not reproduce the accents, code-switching and spontaneous disfluency most relevant to real-world multilingual deployment. The absence of a detected gradient should therefore not be interpreted as equivalence and requires confirmation in recorded clinical speech. Language barriers are independently associated with adverse events and incomplete communication [18,19], so poorer transcription quality remains consequential.

A second equity issue concerns evaluation infrastructure itself. Sixteen languages could not be included in the primary clinical-risk analysis because reliable neural speech synthesis was unavailable; 15 of the 16 were in Joshi classes 0 to 2, with a median class of 1 compared with 3 in the included set (Table S7). Languages most in need of careful evaluation are also among those for which suitable evaluation infrastructure is least available.

### 5.5 Limitations

Limitations define the scope of inference. Synthetic speech strengthened reference control but did not reproduce accents, speaker variability, code-switching or spontaneous disfluency. Risk inference covered 77 of 99 languages and one production transcriber; other systems may have different error distributions. Each complexity level used one fixed script, so scenario and complexity effects cannot be separated, and translation from one English source may under-represent idiomatic or culturally specific language. Serious events were sparse: reliability of the serious-event rate was estimated at 0.740 for a typical language with 30 observations, and power was approximately 44% for a true ρ=0.27. Smaller latent associations and resource-risk gradients therefore remain possible. Three independent large language model raters showed internal consistency but do not constitute a clinical reference standard; shared model dependencies may correlate judgements. A blinded human-validation study of related documentation-error judges found judge-clinician agreement comparable with clinician-clinician agreement [34], but it does not directly calibrate this multilingual instrument. Replication across systems, recorded speech and clinician panels, with calibration to consequential outcomes, is required.

## 6. Conclusion

Across this controlled multilingual corpus, word error rate and related frequency metrics did not reliably track the sparse severe tail of clinically consequential transcription errors. They remain appropriate measures of transcription quality, while context-aware assessment of consequence provides complementary safety information. The observed accuracy gradient without a detectable resource-related safety gradient remains a hypothesis for confirmation in recorded clinical audio.

## CRediT authorship contribution statement

Henry Bergman: Conceptualization, Methodology, Formal analysis, Investigation, Data curation, Writing - original draft, Supervision, Project administration.

Vivian Liu: Methodology, Validation, Writing - review & editing.

Ben Austin: Methodology, Validation, Writing - review & editing.

Rohan Sangera: Methodology, Software, Investigation, Data curation, Validation, Visualization, Writing - review & editing.

## Ethics statement

No human participants and no patient data were involved. All audio was synthesised from author-written scripts containing no patient-identifiable information, so ethical approval was not required.

## Data availability

Data supporting the findings of this study are reported in the article and accompanying Supplementary Materials. The Supplementary Materials provide the methodological specifications necessary to apply the evaluation framework and the variable schema for the frozen per-observation analysis dataset. The underlying evaluation datasets, source materials, and analysis or evaluation code may contain proprietary or commercially sensitive information and are not publicly available. Enquiries regarding access to additional materials may be directed to the corresponding author and will be considered subject to applicable confidentiality, intellectual-property, data-governance, and commercial requirements.

