## Supplementary file for "Beyond word error rate: clinical risk as the necessary standard for ambient AI scribe evaluation: evidence from 77 global languages"

#### Contents


| Item | Title |
|---|---|
| File S1 | Three-rater context-aware clinical-risk scoring specification (rater instructions) |
| File S2 | Method handover: pipeline, orchestration and reproduction notes |
| Analysis A1 | Sensitivity of the language-level correlation to outcome sparsity |
| Table S1 | Per-language transcription results: verbatim and semantic WER, engine, sample size, band and publication tier (99 languages) |
| Table S2 | Occurrence-weighted risk-tier distribution |
| Table S3 | Per-language clinical-risk profile: tier counts, CRITICAL+HIGH percentage, risk density and medical-entity WER (77 languages) |
| Table S4 | Equity covariates by language: GDP per capita, first-language speakers, resource class (74 languages) |
| Table S5 | Consensus dangerous-error register: all CRITICAL patterns with context |
| Table S6 | Consensus dangerous-error register: all HIGH patterns |
| Table S7 | Unspaced-script extension and excluded languages |
| Table S8 | Resource main effects re-centred at in-range complexity values |
| Figure S1 | Occurrence-weighted Severity x Likelihood matrix |
| Figure S2 | Median WER by language and consultation complexity |
| Dataset S1 | Variable schema for the frozen per-observation analysis dataset (28 variables) |

**File S1: Three-rater context-aware clinical-risk scoring specification**

This is the complete rater instruction set, released so that the instrument can be reapplied to any speech-recognition or text-generation error set scored against a ground-truth reference, without reference to the authors.

#### S1.1 What is scored

Each batch is a list of distinct error patterns extracted from the aligned reference and hypothesis. Each pattern carries the reference token or tokens, the hypothesis token or tokens (empty for a deletion), the language, an occurrence count, and worked examples with surrounding context:

```
{ "error_id": "SUB:copd→cop d",
  "type": "substitution" | "deletion",
  "ref":  "<reference token(s)>",
  "hyp":  "<hypothesis token(s), empty for deletion>",
  "lang": "<code>", "count": <occurrences>,
  "examples": [ { "transcript_id", "noise", "lang",
                  "ref_context": "±110 chars around the error in the reference",
                  "hyp_context": "±110 chars around the same point in the hypothesis" } ] }
```

The reference is what was said; the hypothesis is what the transcriber produced. Raters judge the clinical risk of the transcription error in context, reading both context windows. Judging the token in isolation is the single most common way to get this wrong.

#### S1.2 Severity scale (1 to 5)

Clinical consequence if the error reaches the note and is believed.

| Score | Definition |
|---|---|
| 1 | Negligible. Cosmetic; no clinical meaning. |
| 2 | Minor. |
| 3 | Moderate. Could mislead, but would likely be caught. |
| 4 | Major. Could change management. |
| 5 | Catastrophic. Could cause serious harm or death, for example drug, dose, laterality, negation or diagnosis flips. |

#### S1.3 Likelihood scale (1 to 5)

Probability that the error both survives review and propagates to harm.

| Score | Definition |
|---|---|
| 1 | Very unlikely. Obvious nonsense, self-evidently wrong, or the value reappears correctly elsewhere in the note. |
| 2 | Unlikely. |
| 3 | Possible. |
| 4 | Likely. |
| 5 | Very likely. Fluent, plausible and undetectable on ordinary reading. |

**Tier = Severity x Likelihood:** CRITICAL 16 to 25; HIGH 10 to 15; MEDIUM 5 to 9; LOW 1 to 4.

#### S1.4 The four context-aware judging rules

1. **Judge with the surrounding text, never the token alone.** A deleted spoken number that reappears as a digit elsewhere in the note scores likelihood 1. A garbled token that reads as obvious nonsense, which no clinician would believe, scores likelihood 1.
2. **Score high-severity classes up, even when subtle.** Negation flips (resectable to unresectable; no to now; taken to stopped), drug swaps (FOLFOX to FOLFIRINOX; clopidogrel to thyroxine), dose, unit and laterality changes, and diagnosis substitutions (NSTEMI to STEMI).
3. **Score low classes down.** Register, formatting and spelling variants; function words; and number-format differences whose value survives elsewhere.

4. **Default sceptical.** LOW or MEDIUM unless the in-context reading is genuinely dangerous. Scoring words in isolation over-estimated the CRITICAL and HIGH share roughly threefold in development; this rule is the correction for that.

#### S1.5 Pre-scoring exclusions

Raw word-error lists are dominated by differences that are not transcription errors. The following are removed per language before scoring, or the risk denominator is swamped by linguistic noise and the resulting density is meaningless: number-format differences where the value matches; register, inflection, synonym and spelling variants of the same lexeme, which matter particularly for diglossic and morphologically rich languages; code-switching where a native-script transliteration substitutes for the equivalent Latin-script clinical term; and trivial function words and pure punctuation.

**Anything touching a drug, test, numeric value, dose, anatomical site, diagnosis or negation is never filtered as equivalent and is always retained as a genuine error.**

#### S1.6 Rater output and independence

Each of three raters writes one object per input pattern, with the error identifier copied verbatim so the aggregator can join on it:

```
[ { "error_id": "<copied exactly>", "severity": <1-5>,
    "likelihood": <1-5>, "rationale": "<one clause, in context>" }, ... ]
```

Independence means three separate raters scoring fresh from the rubric without sight of one another's scores. In this study, the three raters were instantiated using models from different external providers/model families, none of which was used in the production transcription system under evaluation. One rater scoring three times does not satisfy the specification.

**File S2: Method handover: pipeline, orchestration and reproduction notes**
A self-contained account of how the instrument was operated, sufficient to reproduce the method on a new dataset.

#### S2.1 What the pipeline produces
A risk tier for every distinct error pattern by majority vote of three independent raters; occurrence-weighted risk density, meaning the percentage of error instances falling in the CRITICAL or HIGH tiers, which is the headline quantity; inter-rater agreement as unanimous percentage, majority percentage and Fleiss kappa; and a dangerous-error register listing the consensus CRITICAL and HIGH patterns, which is the actionable output.

The purpose of the method is calibration. Naive scoring of whether an error looks alarming, judged in isolation, over-estimates serious risk roughly threefold. Judging in context, using three raters, and filtering non-errors first together give a defensible and reproducible figure.

#### S2.2 Five-stage pipeline
**Stage 1. Extract and deduplicate patterns with context.** Take the aligned substitutions and deletions for each record, apply the exclusions in File S1.5, and deduplicate into unique patterns keyed on the error itself, retaining an occurrence count and worked examples. The ±110-character context windows are what make in-context judging possible and are non-negotiable. For a deletion, window the hypothesis at the proportional position so the rater sees what the note contains where words were dropped. Unspaced scripts have no word tokens, so patterns are extracted from maximal alignment runs, dropping single-codepoint fragments. Batch into files of roughly 135 to 200 patterns.

**Stage 2. Three independent raters per batch.** Each rater reads the specification and the batch, scores every pattern, and writes its own output file. Independence is enforced by using three separate rater processes from different external model providers/families with no shared context and no access to one another's scores.

**Stage 3. Consensus and agreement.** Tier is the majority vote of the three raters; ties resolve to the tier of the median Severity x Likelihood product. Report the unanimous percentage, the majority percentage and Fleiss kappa together.

**Stage 4. Aggregate to risk density.** Weight each scored pattern's consensus tier by its observed occurrence count. Sampled singleton patterns therefore contribute one occurrence each to the scored-set denominator. No inverse-probability reweighting is applied to represent unsampled singleton patterns; occurrence-weighted estimates describe the scored error set. Report per-language risk density as CRITICAL and HIGH occurrences per 1,000 reference words, the occurrence-weighted Severity x Likelihood matrix, and the dangerous-error register

**Stage 5. Interpret honestly.** Treat proportions and rank orderings as robust and absolute CRITICAL counts as indicative, since per-batch rater calibration varies. State the synthetic-validity caveat where audio is synthesised. Look for patterns in the register, which are more actionable than the counts.

#### S2.3 Scope decision on rare patterns
Scoring every single-occurrence pattern is expensive and low yield, while recurring patterns capture the systematic errors and roughly 80% or more of instances. Frequency filtering is nevertheless inappropriate when hunting dangerous errors because a rare drug swap may be clinically important. The implemented compromise was to score all patterns occurring at least twice plus a stratified sample of singleton patterns. Sampled singletons enter the occurrence-weighted summaries with count 1 and are not up-weighted to represent unsampled singletons; the reported risk distributions are therefore scored-set estimates rather than estimates for the full extracted singleton population. Stratum-specific singleton sampling fractions were not available in the supplied analysis record and are not inferred here.

#### S2.4 Known pitfalls

**Filter non-errors first, and do not over-filter clinical content.** Without the exclusions the density is meaningless; with over-aggressive exclusions genuine clinical errors disappear.

**Context windows flip most alarming deletions to LOW.** In the majority of cases the deleted value reappears elsewhere in the note.

**Use three independent raters.** One rater scoring three times does not produce independent judgements.

**Report the unanimous percentage alongside kappa.** Under a heavily skewed marginal, chance-corrected agreement is deflated even when raw agreement is high. Here kappa was 0.358, conventionally read as fair, against 93% unanimous agreement [1].

**Expect a small number of patterns to recur 20 to 30 times.** One systematically mis-transcribed stock sentence will correctly dominate the occurrence-weighted CRITICAL count, and this should be stated rather than smoothed away.

**Analysis A1: Sensitivity of the language-level correlation to outcome sparsity**

The primary language-level outcome was the proportion of observations containing at least one CRITICAL or HIGH error. A beta-binomial model fitted to the per-language counts (282 events among 5,308 observations overall) separated between-language variation from binomial sampling noise. The fitted mean language-level rate was 0.043 with a between-language standard deviation of 0.060. Estimated reliability of a language's observed rate was 0.740 at the median sample size (n=30), 0.766 at the mean per-language sample size and 0.996 for English. Thus, typical language-level outcomes contain material sampling error that can attenuate rank correlations toward zero.

Simulation under the realised design (77 languages, median 30 observations per language; pooled observation-level serious-event rate 5.31%) estimated approximately 44% power to detect a true Spearman correlation of 0.27. Approximately 80% power was reached at a true correlation of 0.40. These analyses qualify rather than overturn the language-level result: the observed data do not support a large positive association, but smaller or modest latent associations cannot be excluded solely from the non-significant correlations. Precision of the observation-level regression estimates is represented separately by their confidence intervals.

### Table S1: Per-language transcription results

Reported on the best available neural voice per language where one exists. Verbatim WER includes number-format and code-switch corrections; semantic WER additionally allows register, inflection, synonym and spelling variants, which matters for diglossic languages. Unspaced scripts use text canonicalisation and character error rate. Robotic denotes single-speaker non-neural synthesis only, which carries the synthetic-validity caveat and was excluded from risk inference.

| # | Language | Code | Engine | n | Verbatim WER | Semantic WER | Band | Tier |
|---|---|---|---|---|---|---|---|---|
| 1 | English | en | edge+piper | 705 | 0.009 | 0.009 | GREEN | PUBLISH |
| 2 | Indonesian | id | edge | 30 | 0.013 | 0.013 | GREEN | PUBLISH |
| 3 | Vietnamese | vi | edge | 30 | 0.019 | 0.019 | GREEN | PUBLISH |
| 4 | Malay | ms | edge | 30 | 0.020 | 0.020 | GREEN | PUBLISH |
| 5 | Portuguese | pt | edge | 60 | 0.029 | 0.029 | GREEN | PUBLISH |
| 6 | Ukrainian | uk | edge | 30 | 0.030 | 0.030 | GREEN | PUBLISH |
| 7 | Macedonian | mk | edge | 30 | 0.035 | 0.035 | GREEN | PUBLISH |
| 8 | Russian | ru | edge | 30 | 0.035 | 0.035 | GREEN | PUBLISH |
| 9 | Turkish | tr | edge | 30 | 0.035 | 0.035 | GREEN | PUBLISH |
| 10 | French | fr | edge+piper | 135 | 0.036 | 0.036 | GREEN | PUBLISH |
| 11 | Swahili | sw | edge | 30 | 0.036 | 0.036 | GREEN | PUBLISH |
| 12 | Swedish | sv | edge | 30 | 0.038 | 0.038 | GREEN | PUBLISH |
| 13 | Spanish | es | edge | 45 | 0.038 | 0.038 | GREEN | PUBLISH |
| 14 | Norwegian | nb | edge+piper | 30 | 0.042 | 0.042 | GREEN | PUBLISH |
| 15 | Danish | da | edge | 30 | 0.045 | 0.045 | GREEN | PUBLISH |
| 16 | Bulgarian | bg | edge | 30 | 0.045 | 0.045 | GREEN | PUBLISH |
| 17 | Italian | it | edge | 60 | 0.047 | 0.047 | GREEN | PUBLISH |
| 18 | Albanian | sq | edge | 30 | 0.048 | 0.048 | GREEN | PUBLISH |
| 19 | Filipino | fil | edge | 30 | 0.048 | 0.048 | GREEN | PUBLISH |
| 20 | Cebuano | ceb | elevenlabs+mms | 30 | 0.079 | 0.050 | GREEN | PUBLISH |
| 21 | Serbian | sr | edge | 30 | 0.050 | 0.050 | GREEN | PUBLISH |
| 22 | Belarusian | be | elevenlabs | 30 | 0.050 | 0.050 | GREEN | PUBLISH |
| 23 | Bosnian | bs | edge | 30 | 0.050 | 0.050 | GREEN | PUBLISH |
| 24 | Persian | fa | edge | 30 | 0.051 | 0.051 | GREEN | PUBLISH |
| 25 | Armenian | hy | elevenlabs | 30 | 0.053 | 0.053 | GREEN | PUBLISH |
| 26 | Finnish | fi | edge | 30 | 0.057 | 0.057 | GREEN | PUBLISH |
| 27 | Catalan | ca | edge | 30 | 0.105 | 0.059 | GREEN | PUBLISH |
| 28 | Chinese | zh | edge | 60 | 0.059 | 0.059 | GREEN | PUBLISH |
| 29 | Dutch | nl | edge | 75 | 0.059 | 0.059 | GREEN | PUBLISH |
| 30 | Icelandic | is | edge+piper | 30 | 0.060 | 0.060 | GREEN | PUBLISH |
| 31 | Assamese | as | elevenlabs+mms | 30 | 0.075 | 0.062 | GREEN | PUBLISH |
| 32 | Gujarati | gu | edge | 30 | 0.062 | 0.062 | GREEN | PUBLISH |
| 33 | Azerbaijani | az | edge | 30 | 0.067 | 0.067 | GREEN | PUBLISH |
| 34 | German | de | edge+piper | 150 | 0.068 | 0.068 | GREEN | PUBLISH |
| 35 | Arabic | ar | edge | 30 | 0.069 | 0.069 | GREEN | PUBLISH |
| 36 | Latvian | lv | edge | 30 | 0.070 | 0.070 | GREEN | PUBLISH |
| 37 | Javanese | jv | edge | 30 | 0.098 | 0.075 | GREEN | PUBLISH |
| 38 | Romanian | ro | edge | 30 | 0.076 | 0.076 | GREEN | PUBLISH |
| 39 | Afrikaans | af | edge | 30 | 0.077 | 0.077 | GREEN | PUBLISH |
| 40 | Bengali | bn | edge | 30 | 0.078 | 0.078 | GREEN | PUBLISH |
| 41 | Georgian | ka | edge | 30 | 0.078 | 0.078 | GREEN | PUBLISH |
| 42 | Hebrew | he | edge | 30 | 0.080 | 0.080 | GREEN | PUBLISH |
| 43 | Lithuanian | lt | edge | 30 | 0.081 | 0.081 | GREEN | PUBLISH |
| 44 | Greek | el | edge | 30 | 0.081 | 0.081 | GREEN | PUBLISH |
| 45 | Welsh | cy | edge+piper | 30 | 0.082 | 0.082 | GREEN | PUBLISH |
| 46 | Kazakh | kk | edge | 30 | 0.084 | 0.084 | GREEN | PUBLISH |
| 47 | Croatian | hr | edge | 30 | 0.084 | 0.084 | GREEN | PUBLISH |
| 48 | Japanese | ja | edge | 30 | 0.084 | 0.084 | GREEN | PUBLISH |
| 49 | Hindi | hi | edge | 30 | 0.087 | 0.087 | GREEN | PUBLISH |
| 50 | Czech | cs | edge | 30 | 0.091 | 0.091 | GREEN | PUBLISH |
| 51 | Urdu | ur | edge | 60 | 0.092 | 0.092 | GREEN | PUBLISH |
| 52 | Nepali | ne | edge | 30 | 0.103 | 0.092 | GREEN | PUBLISH |
| 53 | Slovak | sk | edge | 30 | 0.093 | 0.093 | GREEN | PUBLISH |
| 54 | Telugu | te | edge | 30 | 0.114 | 0.093 | GREEN | PUBLISH |

| # | Language | Code | Engine | n | Verbatim WER | Semantic WER | Band | Tier |
|---|---|---|---|---|---|---|---|---|
| 55 | Korean | ko | edge | 30 | 0.146 | 0.094 | GREEN | PUBLISH |
| 56 | Kannada | kn | edge | 30 | 0.095 | 0.095 | GREEN | PUBLISH |
| 57 | Tamil | ta | edge | 30 | 0.127 | 0.095 | GREEN | PUBLISH |
| 58 | Galician | gl | edge | 30 | 0.097 | 0.097 | GREEN | PUBLISH |
| 59 | Uzbek | uz | edge | 30 | 0.098 | 0.098 | GREEN | PUBLISH |
| 60 | Polish | pl | edge | 30 | 0.120 | 0.099 | GREEN | PUBLISH |
| 61 | Slovenian | sl | edge | 30 | 0.112 | 0.103 | AMBER | PUBLISH |
| 62 | Punjabi | pa | mms | 15 | 0.104 | 0.104 | AMBER | LOW-CONF |
| 63 | Kyrgyz | ky | elevenlabs+mms | 30 | 0.123 | 0.104 | AMBER | PUBLISH |
| 64 | Sundanese | su | edge | 30 | 0.178 | 0.111 | AMBER | PUBLISH |
| 65 | Thai | th | edge | 30 | 0.111 | 0.111 | AMBER | PUBLISH |
| 66 | Marathi | mr | edge | 30 | 0.146 | 0.119 | AMBER | PUBLISH |
| 67 | Estonian | et | edge | 30 | 0.143 | 0.122 | AMBER | PUBLISH |
| 68 | Hungarian | hu | edge+piper | 30 | 0.142 | 0.128 | AMBER | PUBLISH |
| 69 | Zulu | zu | edge | 30 | 0.151 | 0.129 | AMBER | PUBLISH |
| 70 | Khmer | km | edge | 30 | 0.136 | 0.132 | AMBER | PUBLISH |
| 71 | Somali | so | edge | 30 | 0.187 | 0.137 | AMBER | PUBLISH |
| 72 | Malayalam | ml | edge | 30 | 0.174 | 0.147 | AMBER | PUBLISH |
| 73 | Mongolian | mn | edge | 30 | 0.188 | 0.149 | AMBER | PUBLISH |
| 74 | Nyanja | ny | elevenlabs+mms | 30 | 0.181 | 0.158 | AMBER | PUBLISH |
| 75 | Kurdish | ku | piper | 15 | 0.166 | 0.166 | AMBER | LOW-CONF |
| 76 | Hausa | ha | elevenlabs+mms | 30 | 0.186 | 0.169 | AMBER | PUBLISH |
| 77 | Tajik | tg | mms | 15 | 0.176 | 0.176 | AMBER | LOW-CONF |
| 78 | Luxembourgish | lb | elevenlabs+piper | 28 | 0.224 | 0.195 | AMBER | PUBLISH |
| 79 | Pashto | ps | edge | 30 | 0.216 | 0.200 | RED | PUBLISH |
| 80 | Maltese | mt | edge | 30 | 0.232 | 0.210 | RED | PUBLISH |
| 81 | Lao | lo | edge | 30 | 0.229 | 0.226 | RED | PUBLISH |
| 82 | Amharic | am | edge | 30 | 0.270 | 0.239 | RED | PUBLISH |
| 83 | Sindhi | sd | elevenlabs | 30 | 0.265 | 0.239 | RED | PUBLISH |
| 84 | Odia (Oriya) | or | mms | 15 | 0.252 | 0.252 | RED | LOW-CONF |
| 85 | Burmese | my | edge | 30 | 0.316 | 0.292 | RED | PUBLISH |
| 86 | Basque | eu | mms | 15 | 0.315 | 0.315 | RED | LOW-CONF |
| 87 | Irish | ga | edge | 30 | 0.344 | 0.332 | RED | PUBLISH |
| 88 | Haitian Creole | ht | mms | 15 | 0.395 | 0.395 | RED | LOW-CONF |
| 89 | Krio | kri | mms | 15 | 0.397 | 0.397 | RED | LOW-CONF |
| 90 | Sinhala | si | edge | 30 | 0.439 | 0.398 | RED | PUBLISH |
| 91 | Meiteilon | mni | parler | 15 | 0.406 | 0.406 | RED | LOW-CONF |
| 92 | West Frisian | fy | piper | 15 | 0.455 | 0.455 | RED | LOW-CONF |
| 93 | Yoruba | yo | mms | 15 | 0.469 | 0.469 | RED | LOW-CONF |
| 94 | Samoan | sm | mms | 15 | 0.596 | 0.596 | RED | LOW-CONF |
| 95 | Malagasy | mg | mms | 15 | 0.661 | 0.661 | RED | LOW-CONF |
| 96 | Shona | sn | mms | 15 | 0.676 | 0.676 | RED | LOW-CONF |
| 97 | Xhosa | xh | coqui | 15 | 0.714 | 0.714 | RED | LOW-CONF |
| 98 | Sesotho | st | coqui | 15 | 0.762 | 0.762 | RED | LOW-CONF |
| 99 | Dhivehi | dv | mms | 15 | 0.994 | 0.994 | RED | LOW-CONF |

Band: GREEN, AMBER or RED quality banding as applied in release review. Tier: publication readiness classification.

### Table S2. Occurrence-weighted risk-tier distribution

| Tier | Severity x Likelihood | Occurrences | % of total |
|---|---|---|---|
| CRITICAL | 16 to 25 | 60 | 0.1% |
| HIGH | 10 to 15 | 191 | 0.3% |
| MEDIUM | 5 to 9 | 1,239 | 2.1% |
| LOW | 1 to 4 | 58,329 | 97.5% |
| Total | | 59,819 | 100% |

Across 9,110 unique error patterns, each scored by three independent raters with tier assigned by majority vote. CRITICAL and HIGH together account for 251 of 59,819 scored occurrences, or 0.42%. Inter-rater agreement: 93% of patterns scored identically by all three raters, 100% resolved by majority, no pattern with three differing tiers. Fleiss kappa 0.358, which is deflated by the predominance of the LOW category and should be read alongside the unanimity rate.[1,2]

### Table S3. Per-language clinical-risk profile

Consensus tier counts by language across the 77 spaced neural-voiced languages meeting the inclusion criterion. Risk density is CRITICAL and HIGH occurrences per 1,000 reference words. ME-WER is the medical entity error rate over abbreviations, drug names, laboratory terms and proper nouns.

| Language | WER band | CRIT | HIGH | MED | LOW | Unique patterns | % CRIT+HIGH | Risk density | ME-WER |
|---|---|---|---|---|---|---|---|---|---|
| Galician | GREEN | 0 | 1 | 1 | 120 | 122 | 1.9% | 1.22 | 0.159 |
| Portuguese | GREEN | 0 | 1 | 1 | 96 | 98 | 1.5% | 0.62 | 0.167 |
| German | GREEN | 2 | 0 | 2 | 240 | 244 | 1.5% | 0.85 | 0.057 |
| Hausa | RED | 1 | 3 | 7 | 200 | 211 | 1.0% | 0.68 | 0.224 |
| Nyanja (Chichewa) | RED | 0 | 3 | 11 | 266 | 280 | 1.0% | 0.80 | 0.275 |
| Vietnamese | GREEN | 0 | 1 | 0 | 60 | 61 | 0.8% | 0.33 | 0.046 |
| Slovenian | AMBER | 0 | 1 | 0 | 64 | 65 | 0.8% | 0.32 | 0.073 |
| English | GREEN | 4 | 16 | 72 | 531 | 623 | 0.7% | 0.29 | 0.066 |
| Spanish | GREEN | 0 | 1 | 0 | 76 | 77 | 0.6% | 0.27 | 0.065 |
| Albanian | GREEN | 1 | 0 | 0 | 84 | 85 | 0.6% | 0.26 | 0.133 |
| Macedonian | GREEN | 0 | 1 | 0 | 68 | 69 | 0.6% | 0.29 | 0.109 |
| Mongolian | RED | 1 | 0 | 3 | 108 | 112 | 0.6% | 0.31 | 0.303 |
| Serbian | GREEN | 0 | 1 | 0 | 79 | 80 | 0.5% | 0.29 | 0.093 |
| Filipino (Tagalog) | — | 1 | 0 | 1 | 87 | 89 | 0.5% | 0.27 | 0.165 |
| Persian | GREEN | 0 | 1 | 2 | 89 | 92 | 0.5% | 0.27 | 0.222 |
| Gujarati | GREEN | 0 | 1 | 0 | 152 | 153 | 0.3% | 0.29 | 0.094 |
| Icelandic | GREEN | 0 | 1 | 3 | 197 | 201 | 0.2% | 0.15 | 0.234 |
| Cebuano | — | 0 | 1 | 7 | 132 | 140 | 0.2% | 0.08 | 0.168 |
| French | GREEN | 0 | 1 | 3 | 215 | 219 | 0.1% | 0.05 | 0.092 |
| Afrikaans | GREEN | 0 | 0 | 1 | 122 | 123 | 0.0% | 0.00 | 0.191 |
| Amharic | RED | 0 | 0 | 2 | 147 | 149 | 0.0% | 0.00 | 0.174 |
| Arabic | GREEN | 0 | 0 | 0 | 103 | 103 | 0.0% | 0.00 | 0.080 |
| Assamese | AMBER | 0 | 0 | 6 | 146 | 152 | 0.0% | 0.00 | 0.328 |
| Azerbaijani | GREEN | 0 | 0 | 4 | 97 | 101 | 0.0% | 0.00 | 0.194 |
| Belarusian | GREEN | 0 | 0 | 1 | 88 | 89 | 0.0% | 0.00 | 0.050 |
| Bulgarian | GREEN | 0 | 0 | 2 | 86 | 88 | 0.0% | 0.00 | 0.096 |
| Bengali | GREEN | 0 | 0 | 2 | 116 | 118 | 0.0% | 0.00 | 0.176 |
| Bosnian | GREEN | 0 | 0 | 1 | 50 | 51 | 0.0% | 0.00 | 0.081 |
| Catalan | AMBER | 0 | 0 | 2 | 92 | 94 | 0.0% | 0.00 | 0.130 |
| Czech | GREEN | 0 | 0 | 0 | 76 | 76 | 0.0% | 0.00 | 0.026 |
| Welsh | GREEN | 0 | 0 | 5 | 161 | 166 | 0.0% | 0.00 | 0.286 |
| Danish | GREEN | 0 | 0 | 0 | 75 | 75 | 0.0% | 0.00 | 0.101 |
| Greek | GREEN | 0 | 0 | 0 | 101 | 101 | 0.0% | 0.00 | 0.141 |
| Estonian | AMBER | 0 | 0 | 2 | 95 | 97 | 0.0% | 0.00 | 0.128 |
| Finnish | GREEN | 0 | 0 | 3 | 85 | 88 | 0.0% | 0.00 | 0.059 |
| Irish | RED | 0 | 0 | 4 | 133 | 137 | 0.0% | 0.00 | 0.487 |
| Hebrew | GREEN | 0 | 0 | 2 | 120 | 122 | 0.0% | 0.00 | 0.074 |
| Hindi | GREEN | 0 | 0 | 0 | 153 | 153 | 0.0% | 0.00 | 0.037 |
| Croatian | GREEN | 0 | 0 | 0 | 74 | 74 | 0.0% | 0.00 | 0.092 |
| Hungarian | AMBER | 0 | 0 | 4 | 158 | 162 | 0.0% | 0.00 | 0.169 |
| Armenian | GREEN | 0 | 0 | 0 | 71 | 71 | 0.0% | 0.00 | 0.154 |
| Indonesian | GREEN | 0 | 0 | 0 | 47 | 47 | 0.0% | 0.00 | 0.020 |
| Italian | GREEN | 0 | 0 | 3 | 92 | 95 | 0.0% | 0.00 | 0.059 |
| Javanese | GREEN | 0 | 0 | 3 | 67 | 70 | 0.0% | 0.00 | 0.213 |
| Georgian | GREEN | 0 | 0 | 1 | 59 | 60 | 0.0% | 0.00 | 0.181 |
| Kazakh | GREEN | 0 | 0 | 2 | 86 | 88 | 0.0% | 0.00 | 0.448 |
| Kannada | GREEN | 0 | 0 | 0 | 117 | 117 | 0.0% | 0.00 | 0.165 |
| Korean | AMBER | 0 | 0 | 2 | 99 | 101 | 0.0% | 0.00 | 0.148 |
| Kyrgyz | AMBER | 0 | 0 | 3 | 149 | 152 | 0.0% | 0.00 | 0.281 |
| Luxembourgish | RED | 0 | 0 | 4 | 227 | 231 | 0.0% | 0.00 | 0.247 |
| Lithuanian | GREEN | 0 | 0 | 0 | 113 | 113 | 0.0% | 0.00 | 0.139 |
| Latvian | GREEN | 0 | 0 | 0 | 105 | 105 | 0.0% | 0.00 | 0.109 |
| Malayalam | AMBER | 0 | 0 | 1 | 87 | 88 | 0.0% | 0.00 | 0.276 |
| Marathi | AMBER | 0 | 0 | 0 | 95 | 95 | 0.0% | 0.00 | 0.080 |
| Malay | GREEN | 0 | 0 | 0 | 38 | 38 | 0.0% | 0.00 | 0.044 |

| Language | WER band | CRIT | HIGH | MED | LOW | Unique patterns | % CRIT+HIGH | Risk density | ME-WER |
|---|---|---|---|---|---|---|---|---|---|
| Maltese | RED | 0 | 0 | 2 | 95 | 97 | 0.0% | 0.00 | 0.256 |
| Norwegian | GREEN | 0 | 0 | 2 | 63 | 65 | 0.0% | 0.00 | 0.110 |
| Nepali | AMBER | 0 | 0 | 4 | 87 | 91 | 0.0% | 0.00 | 0.106 |
| Dutch | GREEN | 0 | 0 | 2 | 81 | 83 | 0.0% | 0.00 | 0.053 |
| Polish | AMBER | 0 | 0 | 0 | 101 | 101 | 0.0% | 0.00 | 0.056 |
| Pashto | RED | 0 | 0 | 0 | 190 | 190 | 0.0% | 0.00 | 0.232 |
| Romanian | GREEN | 0 | 0 | 1 | 128 | 129 | 0.0% | 0.00 | 0.057 |
| Russian | GREEN | 0 | 0 | 0 | 82 | 82 | 0.0% | 0.00 | 0.063 |
| Sindhi | RED | 0 | 0 | 2 | 174 | 176 | 0.0% | 0.00 | 0.104 |
| Sinhala | RED | 0 | 0 | 0 | 110 | 110 | 0.0% | 0.00 | 0.522 |
| Slovak | GREEN | 0 | 0 | 0 | 80 | 80 | 0.0% | 0.00 | 0.043 |
| Somali | AMBER | 0 | 0 | 2 | 107 | 109 | 0.0% | 0.00 | 0.341 |
| Sundanese | AMBER | 0 | 0 | 0 | 105 | 105 | 0.0% | 0.00 | 0.172 |
| Swedish | GREEN | 0 | 0 | 0 | 90 | 90 | 0.0% | 0.00 | 0.047 |
| Swahili | GREEN | 0 | 0 | 0 | 66 | 66 | 0.0% | 0.00 | 0.072 |
| Tamil | AMBER | 0 | 0 | 0 | 96 | 96 | 0.0% | 0.00 | 0.094 |
| Telugu | AMBER | 0 | 0 | 0 | 86 | 86 | 0.0% | 0.00 | 0.076 |
| Turkish | GREEN | 0 | 0 | 1 | 70 | 71 | 0.0% | 0.00 | 0.052 |
| Ukrainian | GREEN | 0 | 0 | 0 | 70 | 70 | 0.0% | 0.00 | 0.088 |
| Urdu | GREEN | 0 | 0 | 6 | 206 | 212 | 0.0% | 0.00 | 0.088 |
| Uzbek | GREEN | 0 | 0 | 1 | 104 | 105 | 0.0% | 0.00 | 0.205 |
| Zulu | AMBER | 0 | 0 | 4 | 81 | 85 | 0.0% | 0.00 | 0.218 |

Percentages are occurrence-weighted. Languages are ordered by descending CRITICAL and HIGH percentage.

### Table S4: Equity covariates by language

GDP per capita in thousands of US dollars for the dominant first-language country (International Monetary Fund, 2026, nominal) [3]. First-language speakers are in millions (Ethnologue, 26th edition) [4]. Resource class 0 to 5 follows documented availability of language-technology resources [5]. This table contains 74 complete external-covariate records from the 77-language primary risk set; Malayalam (ml), Slovak (sk) and Urdu (ur) are not represented because complete external-covariate rows were unavailable in the supplied equity table. No imputation was applied. Clinical-risk outcomes are reported once, in Table S3, which is the authoritative per-language risk table.

| Language code | WER | Resource class | GDP per capita ($k) | Speakers (m) | GDP source | Speaker source |
|---|---|---|---|---|---|---|
| af | 0.077 | 3 | 7.5 | 7.2 | verified | approx |
| am | 0.239 | 2 | 1.08 | 32.0 | verified | approx |
| ar | 0.069 | 5 | 3.9 | 83.0 | verified | verified |
| as | 0.078 | 1 | 2.8 | 15.0 | verified | approx |
| az | 0.067 | 1 | 7.5 | 23.0 | verified | approx |
| be | 0.050 | 3 | 7.8 | 5.0 | approx | approx |
| bg | 0.045 | 3 | 23.8 | 8.0 | verified | approx |
| bn | 0.078 | 3 | 2.9 | 234.0 | verified | verified |
| bs | 0.050 | 1 | 8.4 | 2.5 | approx | approx |
| ca | 0.059 | 4 | 41.6 | 4.5 | verified | approx |
| ceb | 0.080 | 1 | 4.4 | 20.0 | verified | approx |
| cs | 0.091 | 4 | 39.8 | 10.0 | verified | approx |
| cy | 0.099 | 1 | 49.0 | 0.9 | approx | approx |
| da | 0.045 | 3 | 83.4 | 5.5 | verified | approx |
| de | 0.071 | 5 | 65.3 | 76.0 | verified | verified |
| el | 0.081 | 3 | 29.7 | 13.0 | verified | approx |
| en | 0.017 | 5 | 94.4 | 372.0 | verified | verified |
| es | 0.038 | 5 | 41.6 | 487.0 | verified | verified |
| et | 0.122 | 3 | 37.7 | 1.0 | verified | approx |
| fa | 0.051 | 4 | 3.4 | 70.0 | verified | approx |
| fi | 0.057 | 5 | 60.1 | 5.0 | verified | approx |
| fil | 0.048 | 3 | 4.4 | 45.0 | verified | approx |
| fr | 0.037 | 5 | 52.1 | 75.0 | verified | verified |
| ga | 0.332 | 2 | 140.2 | 1.2 | verified | approx |
| gl | 0.097 | 1 | 41.6 | 2.4 | verified | approx |
| gu | 0.063 | 1 | 2.8 | 58.0 | verified | verified |
| ha | 0.213 | 2 | 1.56 | 58.0 | verified | verified |
| he | 0.080 | 3 | 69.8 | 9.0 | verified | approx |
| hi | 0.087 | 4 | 2.8 | 347.0 | verified | verified |
| hr | 0.084 | 4 | 30.0 | 5.0 | verified | approx |
| hu | 0.144 | 4 | 28.4 | 13.0 | verified | approx |
| hy | 0.053 | 1 | 10.4 | 6.7 | verified | approx |
| id | 0.013 | 3 | 5.4 | 78.0 | verified | verified |
| is | 0.060 | 2 | 110.0 | 0.36 | verified | approx |
| it | 0.047 | 5 | 46.5 | 60.0 | verified | verified |

| | | | | | | |
|---|---|---|---|---|---|---|
| jv | 0.075 | 1 | 5.4 | 82.0 | verified | approx |
| ka | 0.078 | 3 | 11.6 | 3.7 | verified | approx |
| kk | 0.084 | 3 | 17.5 | 13.0 | verified | approx |
| kn | 0.095 | 1 | 2.8 | 44.0 | verified | verified |
| ko | 0.094 | 4 | 37.4 | 82.0 | verified | verified |
| ky | 0.118 | 1 | 2.0 | 5.0 | approx | approx |
| lb | 0.211 | 1 | 158.7 | 0.4 | verified | approx |
| lt | 0.081 | 3 | 36.5 | 3.0 | verified | approx |
| lv | 0.070 | 3 | 28.9 | 1.5 | verified | approx |
| mk | 0.035 | 1 | 12.0 | 1.4 | verified | approx |
| mn | 0.149 | 1 | 7.9 | 5.0 | verified | approx |
| mr | 0.119 | 2 | 2.8 | 83.0 | verified | verified |
| ms | 0.020 | 3 | 12.6 | 33.0 | approx | approx |
| mt | 0.210 | 2 | 53.6 | 0.5 | verified | approx |
| nb | 0.047 | 1 | 105.9 | 5.0 | verified | approx |
| ne | 0.092 | 1 | 1.55 | 16.0 | verified | approx |
| nl | 0.059 | 5 | 79.9 | 25.0 | verified | approx |
| ny | 0.185 | 1 | 0.73 | 14.0 | verified | approx |
| pl | 0.099 | 4 | 31.3 | 40.0 | verified | approx |
| ps | 0.200 | 1 | 0.45 | 50.0 | verified | approx |
| pt | 0.029 | 5 | 35.4 | 252.0 | verified | verified |
| ro | 0.076 | 3 | 25.7 | 24.0 | verified | approx |
| ru | 0.035 | 5 | 18.5 | 133.0 | verified | verified |
| sd | 0.239 | 0 | 1.7 | 25.0 | verified | approx |
| si | 0.398 | 0 | 4.5 | 16.0 | verified | approx |
| sl | 0.103 | 3 | 40.6 | 2.0 | verified | approx |
| so | 0.137 | 1 | 0.81 | 22.0 | verified | approx |
| sq | 0.048 | 1 | 12.5 | 7.5 | verified | approx |
| sr | 0.050 | 4 | 17.3 | 9.0 | verified | approx |
| su | 0.111 | 1 | 5.4 | 42.0 | verified | approx |
| sv | 0.038 | 4 | 70.7 | 10.0 | verified | approx |
| sw | 0.036 | 2 | 1.4 | 18.0 | approx | approx |
| ta | 0.095 | 3 | 2.8 | 79.0 | verified | verified |
| te | 0.093 | 1 | 2.8 | 83.0 | verified | verified |
| tr | 0.035 | 4 | 19.0 | 86.0 | verified | verified |
| uk | 0.030 | 3 | 7.0 | 33.0 | verified | approx |
| uz | 0.098 | 3 | 4.7 | 34.0 | verified | approx |
| vi | 0.019 | 4 | 5.1 | 86.0 | verified | verified |
| zu | 0.129 | 2 | 7.5 | 12.0 | verified | approx |

Source labels record whether each covariate was verified against the primary source or entered as a documented approximation. Hindi illustrates why GDP per capita and resource class should be treated as separate covariates: low GDP per capita, approximately 347 million first-language speakers and comparatively low WER.

### Table S5: Consensus dangerous-error register: CRITICAL patterns

Nine consensus CRITICAL patterns identified in the scored register are shown with the hypothesis context in which the error arose. Per-pattern occurrence counts are omitted here; aggregate CRITICAL and HIGH occurrence totals in Table S2 are the authoritative quantitative denominator. Together with the 25 HIGH patterns in Table S6, the register contains 34 distinct serious-error patterns for mechanistic inspection.

| Language | Count | Error pattern | Context in the transcribed note |
|---|---|---|---|
| de | 29 | SUB:eingenommen→abgesetzt | … rechte Hepatektomie – anstreben. |
| en | 15 | SUB:unchanged→changed | … ia at present. Retinopathy screening from last month was reported as background retinopat… |
| sq | 2 | SUB:folfox→folfiri | … në klopidogrel. |
| de | 2 | SUB:gliclazid→glipizid | … ahr. |
| en | 2 | SUB:bevacizumab→cetuximab | … |
| ha | 2 | SUB:clopidogrel→thyroxine | … ya na baya ya haɗa da type 2 diabetes kan metformin, hawan jini kan ramipril da amlodipin… |
| mn | 2 | SUB:рамиприл→амитриптилин | … он ALT хэвийн, креатинин клиренс 68 байна. |
| en | 2 | SUB:ramipril→omeprazole | … , creatinine unclear and eGFR 62. |
| en | 2 | SUB:the clopidogrel→that bevacizumab | … we would aim for a synchronous or staged resection, sigmoid colectomy, and right hepatect… |

SUB denotes a substitution and DEL a deletion. The register is presented to characterise clinically important error mechanisms; quantitative inference uses the aggregate frozen totals in Table S2 rather than sums of individual register rows.

### Table S6. Consensus dangerous-error register: HIGH patterns

| Language | Count | Error pattern |
|---|---|---|
| en | 49 | SUB:hepatectomy→hemicolectomy |
| en | 25 | SUB:gcsf→renal |
| en | 9 | SUB:tia→dvt |
| gl | 9 | DEL:e un |
| pt | 9 | DEL:dois gramas de |
| en | 8 | SUB:bevacizumab→panitumumab |
| en | 4 | SUB:rate→warfarin |
| vi | 3 | SUB:folfox→folfirinox |
| en | 2 | SUB:nstemi→cabg |
| ha | 2 | SUB:lita biyu→room air |
| ha | 2 | SUB:colectomy→cholecystectomy |
| ha | 2 | SUB:clopidogrel→tirodorel |
| en | 2 | SUB:increased→decreased |
| tr | 2 | SUB:nstemi→stemi |
| sr | 2 | SUB:nstemi→jem |
| fil | 2 | SUB:gliclazide→glipizide |
| mk | 2 | SUB:nstemi→инфаркт |
| is | 2 | SUB:blóðþurrðarhjartasjúkdóm→blóðþrýstingssjúkdóm |
| en | 2 | SUB:ctcae→cabg |
| sl | 2 | SUB:nstemi pri dvaindvajsetih→stemi atrijska fibrilacija |
| en | 2 | SUB:vte→reassessment |
| fr | 2 | SUB:egfr→tsh |
| gu | 2 | SUB:જોતાં gcsf→જોતા gfr |
| en | 2 | SUB:ipap sixteen→iv salbutamol |
| ceb | 1 | SUB:ramipril→metoprolol |

The register is dominated by two mechanisms. Medication-name confusions (clopidogrel to thyroxine, ramipril to amitriptyline or omeprazole, bevacizumab to cetuximab or panitumumab, gliclazide to glipizide, FOLFOX to FOLFIRI or FOLFIRINOX) reflect the look-alike and sound-alike problem documented in medication safety [6]. Negation and clinical-status flips (eingenommen to abgesetzt, meaning taken to discontinued; unchanged to changed; increased to decreased; NSTEMI to STEMI) reflect the assertion-status problem long recognised in clinical language processing [7]. Both classes are dangerous precisely because the output is fluent and plausible and would not present to a reviewing clinician as an error.

### Table S7: Unspaced-script extension and excluded languages

Burmese, Thai, Khmer and Lao use character-level error computation, so genuine errors were extracted as word or morpheme-level alignment-run substitutions with sub-character fragments dropped, then scored separately by the same three-rater procedure.

| Measure | Value |
|---|---|
| Languages | 4 (Burmese, Thai, Khmer, Lao) |
| Unique patterns scored | 172 |
| CRITICAL | 0 |
| HIGH | 0 to 1 consensus |
| MEDIUM | 1 |
| Combined coverage with spaced languages | 81 languages, 0.3% to 0.4% CRITICAL or HIGH |

The single MEDIUM pattern was a Lao home-oxygen value rendered as 4 L in place of 2 L, with the correct value restated adjacently in the note. The low risk-density pattern therefore holds for these languages, arguably more strongly than for the spaced set.

### Table S8: Resource main effects re-centred at in-range complexity values

| Complexity value at which the contrast is evaluated | WER, low vs high resource | CRITICAL/HIGH, low vs high resource |
|---|---|---|
| Complexity = 0 (uncentred) | β = +0.068 (95% CI +0.032 to +0.104), p < 0.001 | OR 3.15 (95% CI 0.32 to 31.06), p = 0.33 |
| Complexity = 3 (level at which dangerous errors first appear) | β = +0.078 (95% CI +0.045 to +0.111), p < 0.0001 | OR 1.21 (95% CI 0.43 to 3.43), p = 0.72 |
| Complexity = 3.00 (sample mean) | β = +0.078 (95% CI +0.045 to +0.111), p < 0.0001 | OR 1.21 (95% CI 0.43 to 3.43), p = 0.72 |

Centring complexity changes only the intercept and resource main effects; the complexity slope and resource-by-complexity interaction are identical to the uncentred models in main-text Table 1. Complexity was observed only at levels 1 to 5, so the uncentred resource contrast at complexity=0 is an extrapolation beyond the observed range and into a region in which no CRITICAL/HIGH error occurred. The level-3 contrast is therefore the clinically interpretable in-range estimate.

### Figure S1: Occurrence-weighted Severity x Likelihood matrix

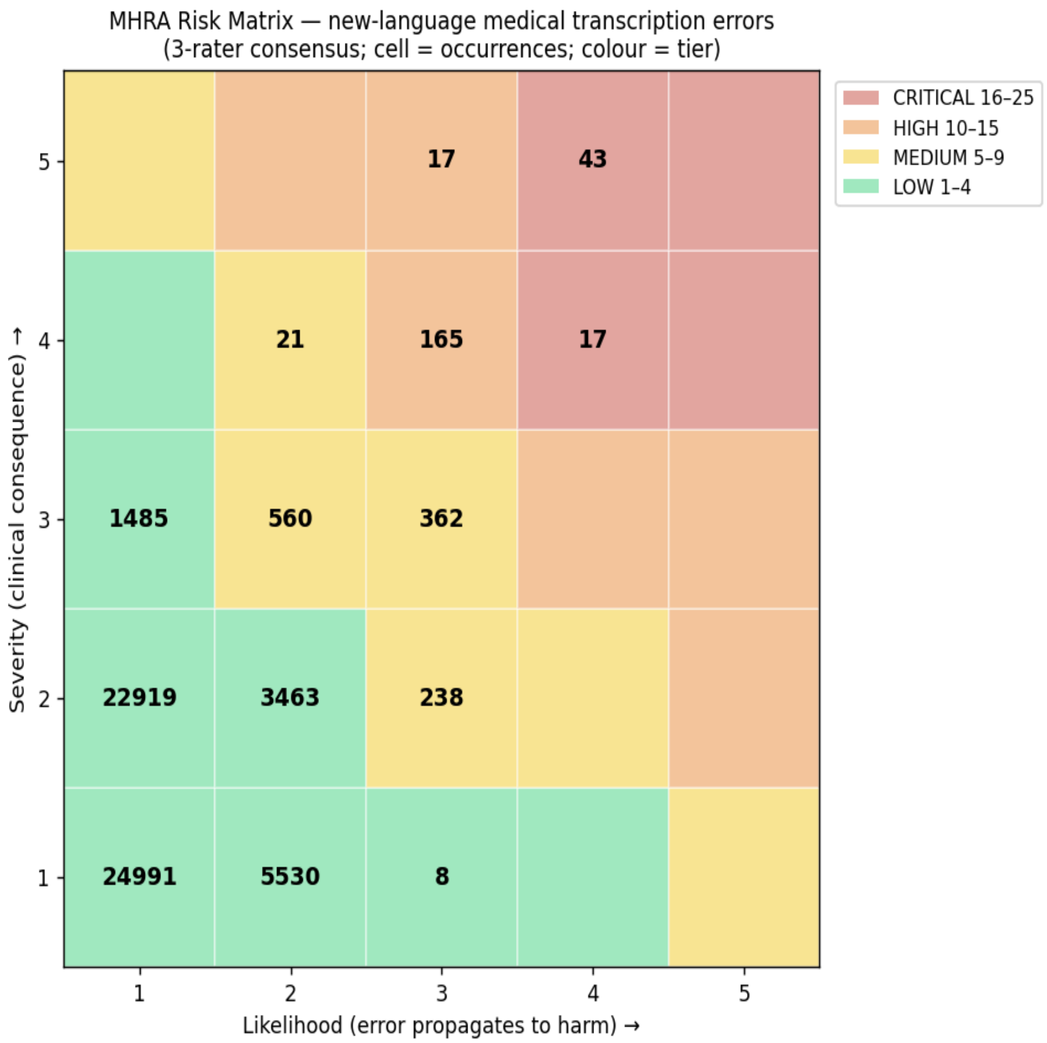


**Figure S1: Occurrence-weighted Severity x Likelihood matrix, three-rater consensus.**

Cell values are numbers of scored error occurrences; colour indicates consequence tier. Occurrences mass in the low-severity, low-likelihood corner. The CRITICAL and HIGH tiers together are reached by 251 of 59,819 occurrences (60 CRITICAL, 191 HIGH; Table S3). Cells containing fewer than eight occurrences are shaded but not labelled, so the annotated values do not sum to the tier totals; the authoritative counts are those in Table S2. Tier boundaries are study-specific; the regulatory and clinical-safety context is described in main-text section 3.7.

### Figure S2: Median WER by language and consultation complexity

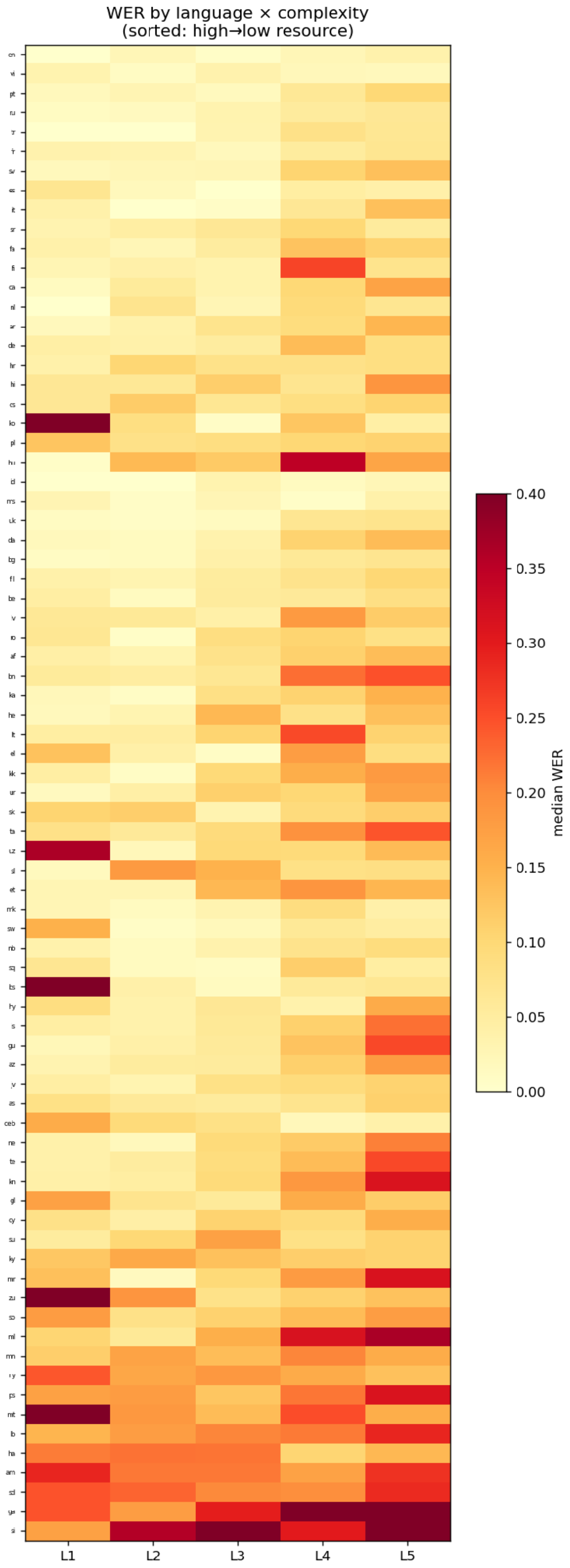
WER by language × complexity
(sorted: high→low resource)
L1
L2
L3
L4
L5
median WER
0.40
0.35
0.30
0.25
0.20
0.15
0.10
0.05
0.00

**Figure S2: Median WER by language and consultation complexity.**

Languages are ordered from high to low resource. WER darkens toward the lower-resource languages and the higher-complexity columns. This is the accuracy gap that, per the main-text figures, was not accompanied by a statistically detectable resource-related safety gradient.

#### Dataset S1: Master per-observation analysis dataset

The frozen per-observation analysis dataset contains 5,758 rows, one per observation crossing language, complexity level, acoustic condition and voice. Dataset S1 provides the variable schema for the 28-variable frozen analysis snapshot used in the reported models. The underlying dataset is not included in the supplementary material, consistent with the Data availability statement in the main manuscript.

| Variable group | Variables |
|---|---|
| Identification | lang, language, transcript_id |
| Design factors | complexity (1 to 5), noise (clean, clinic, ward), voice, engine, neural |
| Language covariates | joshi_class, resource_level, family |
| Frequency metrics | wer, wer_corrected, wer_semantic, me_wer, bleu, chrf, rougeL, bertscore_f1 |
| Error and risk counts | ref_words, n_genuine, error_density, n_crit_high, ch_density, has_crit_high |
| Risk aggregates | risk_sumSL, risk_density_sumSL, risk_assessed |

Three WER variants are supplied because the correction applied changes the value materially in morphologically rich and diglossic languages: wer is verbatim; wer_corrected adds number-format and code-switch equivalence; wer_semantic additionally allows register, inflection, synonym and spelling variants. The main analysis uses wer_semantic. risk_sumSL is the severity-weighted sum reported in the main text as a deliberate contrast, and reduces to an error count for the reason given there.
